# Complement-dependent virion lysis mediated by dengue-Zika virus cross-reactive antibodies correlates with protection from severe dengue disease

**DOI:** 10.1101/2024.06.03.24308395

**Authors:** Antonio G. Dias, Elias Duarte, Jose Victor Zambrana, Jaime A. Cardona-Ospina, Sandra Bos, Vicky Roy, Guillermina Kuan, Angel Balmaseda, Galit Alter, Eva Harris

**Author notes:** These authors contributed equally to this work (Antonio G. Dias, Elias Duarte).

## Abstract

Primary infection with one of four dengue virus serotypes (DENV1-4) may generate antibodies that protect or enhance subsequent secondary heterotypic infections. However, the characteristics of heterotypic cross-reactive antibodies associated with protection from symptomatic infection and severe disease are not well-defined. We selected plasma samples collected before a secondary DENV heterotypic infection that was classified either as dengue fever (DF, n = 31) or dengue hemorrhagic fever/dengue shock syndrome (DHF/DSS, n = 33) from our longstanding pediatric cohort in Nicaragua. We screened various antibody properties to determine the features correlated with protection from DHF/DSS. Protection was associated with high levels of binding of various antibody isotypes, IgG subclasses and effector functions, including antibody-dependent complement deposition, ADCD. Although the samples were derived from DENV-exposed, Zika virus (ZIKV)-naïve individuals, the protective ADCD association was stronger when assays were conducted with recombinant ZIKV antigens. Further, we showed that a complement-mediated virion lysis (virolysis) assay conducted with ZIKV virions was strongly associated with protection, a finding reproduced in an independent sample set collected prior to secondary heterotypic inapparent versus symptomatic DENV infection. Virolysis was the main antibody feature correlated with protection from DHF/DSS and severe symptoms, such as thrombocytopenia, hemorrhagic manifestations, and plasma leakage. Hence, anti-DENV antibodies that cross-react with ZIKV, target virion-associated epitopes, and mediate complement-dependent virolysis are correlated with protection from secondary symptomatic DENV infection and DHF/DSS. These findings may support the rational design and evaluation of dengue vaccines and development of therapeutics.

**One Sentence Summary:** Complement-dependent virolysis mediated by a subset of ZIKV-cross-reactive antibodies protects from symptomatic DENV infection and severe disease.

## INTRODUCTION

The *Flavivirus* genus within the *Flaviviridae* family comprises various medically important mosquito-borne viruses, including dengue viruses (DENV), Zika virus (ZIKV), yellow fever virus (YFV), and West Nile virus (WNV) (*1, 2*). These viruses have caused numerous outbreaks and epidemics throughout history and continue to expand to new areas of the globe (*2*). Approximately 3.9 billion people worldwide live in areas at risk for contracting at least one of the four DENV serotypes (DENV1-4), with an estimated 100 million dengue cases per year (*3*). Symptomatic infections can vary from classical dengue fever (DF) to more severe dengue hemorrhagic fever/shock syndrome (DHF/DSS) (*4*). A primary infection with one DENV serotype generates antibodies that generally protect the host from disease from future homotypic infection with that serotype but may be either protective or enhancing upon subsequent heterotypic infection with a distinct serotype (*5, 6*). The latter phenomenon has been attributed in part to Antibody-Dependent Enhancement (ADE), representing the ability of non-neutralizing or poorly neutralizing cross-reactive antibodies to enhance infection of Fc gamma receptor (FcγR)-bearing myeloid cells (*7–9*). Other potential mechanisms of disease enhancement include high-affinity binding of afucosylated antibodies to the activating FcγRIIIA receptor (*10, 11*). We previously found that individuals with a low-to-intermediate level of cross-reactive binding antibodies were at risk of severe disease during a subsequent DENV infection (*5*). Furthermore, infections by antigenically related flaviviruses can generate cross-reactive antibodies that can be associated with positive and negative outcomes of subsequent heterologous virus infection. For example, we and others have observed that prior DENV infection can be protective of future Zika disease (*12, 13*), while prior ZIKV infection is associated with increased risk of future dengue disease in a serotype-dependent manner (*14, 15*). Hence, by mechanisms that are not clear, DENV and ZIKV infections can generate antibodies that affect the outcome of future DENV and ZIKV infections.

While antibody titers are an important quantitative marker that often correlate with viral infection and disease outcome (*16*), assessing antibody quality (i.e., repertoire and functionality) in DENV infections has proven to be complex. A classical method to assess antibody quality is by conducting neutralization assays. However, measuring neutralizing antibody (nAb) titers against all four DENV serotypes has not always correlated with protection in natural infection studies and vaccine trials. Further, high variability is observed in neutralization assays across laboratories that has been attributed to DENV serotype, virion maturation state, cell substrate, types of antibodies being measured (total versus type-specific versus cross-reactive), and presence or absence of complement, among others (*17–20*). Type-specific DENV nAbs have shown promising results as a correlate of protection, with the limitation that this method often requires depletion of cross-reactive antibodies and larger sample volumes (*19, 21–23*). Therefore, conducting neutralization assays can be challenging and may not always be sufficient to predict the outcome of DENV infections. Our previous study assessing the biophysical and functional properties of the Fc region of anti-DENV antibodies revealed multiple potential biomarkers correlated with protection from a subsequent symptomatic secondary DENV3 infection (*24*). Here, we systematically analyzed multiple antibody properties and found that complement-mediated virion lysis (virolysis) is correlated with protection from severe dengue disease. We found that these protective, virolysis-mediating, anti-DENV antibodies are a specific subgroup of cross-reactive antibodies that bind epitopes present in ZIKV antigens and virions.

## RESULTS

### Pre-existing iELISA and neutralizating antibody titers are not significantly correlated with protection from severe dengue disease

To identify biomarkers that correlate with the outcome of DENV infection and disease, one must analyze pre-secondary infection samples, i.e., samples collected several months or years after an initial primary or multiple prior infection(s) or vaccination, but before a subsequent natural secondary DENV infection. The term ‘secondary infection’ refers to any second, third or fourth subsequent infection. We selected pre-secondary plasma samples from our long-term pediatric cohort in Managua, Nicaragua, to investigate biomarkers of protection correlated with the disease outcomes of subsequent heterotypic DENV infections (*25, 26*). Previously, we had compared individuals with subsequent secondary heterotypic DENV3 infections that were either inapparent or symptomatic (*24*). In the present study, we compared pre-infection samples from individuals with subsequent secondary heterotypic symptomatic infections that were classified as either DF (n=31) or DHF/DSS (n=33) (Fig. 1A). We used pre-secondary infection plasma samples collected between 2005 to 2019 before a secondary heterotypic DENV1 (n=6), DENV2 (n=25), or DENV3 (n=33) infection (Table 1).

**Fig. 1.**
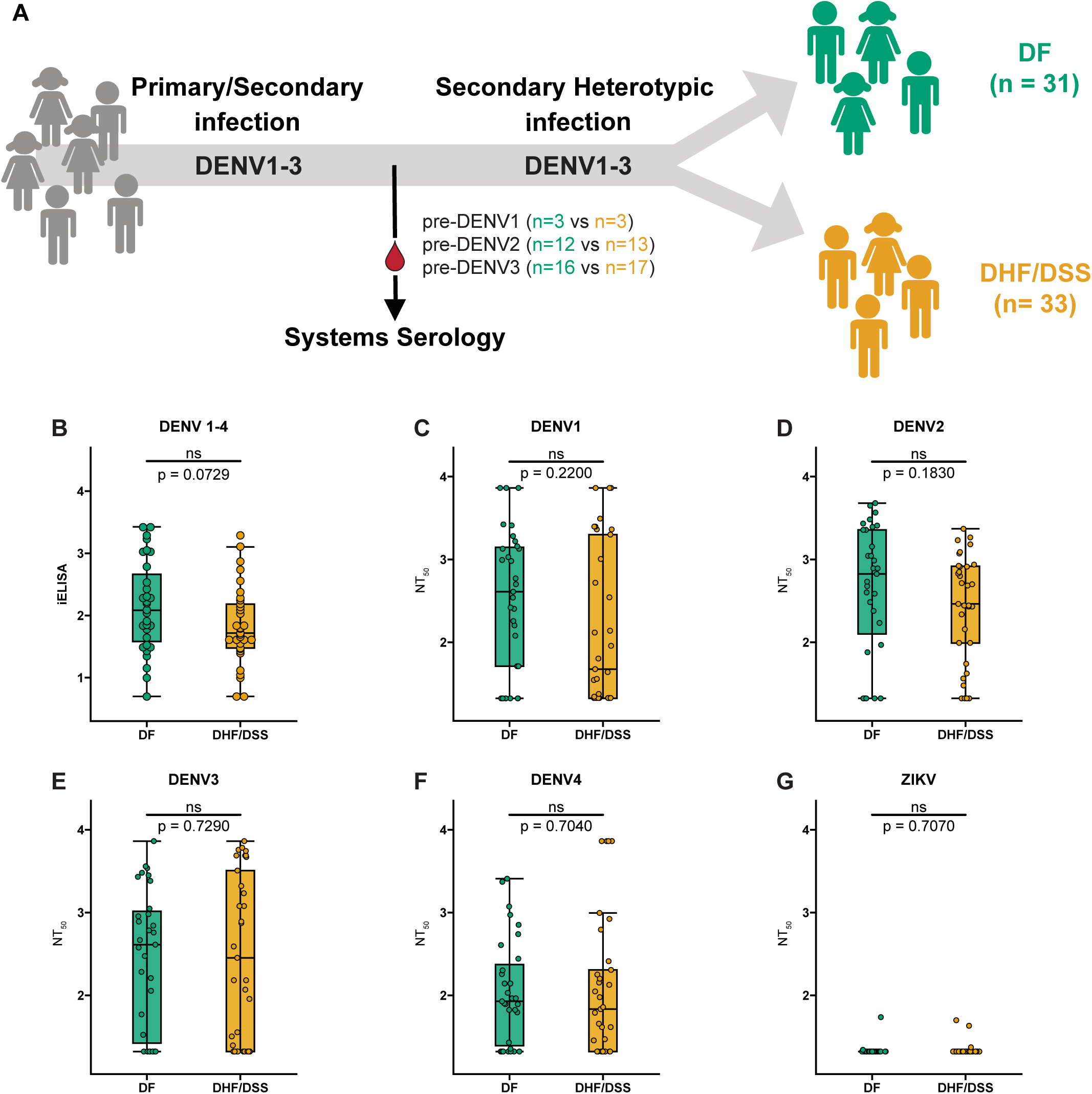
Binding and neutralization titers of pre-infection antibodies are not significantly associated with protection from subsequent severe dengue disease. (A) Plasma samples collected prior to a secondary heterotypic infection resulting in non-severe dengue (DF; green) or severe dengue disease (DHF/DSS; gold) were selected for this systems serology study. (C) Inhibition enzyme-linked immunosorbent assay (iELISA) binding antibody and (C-G) neutralizing antibody titer (NT_50_) of pre-DF and pre-DHF/DSS groups measured against DENV1-4 and ZIKV by FRNT on Vero cells with mature virions. Shown are median NT_50_ (middle line), 25^th^ to 75^th^ percentile (box), and 5^th^ to 95^th^ percentile (whiskers) as well as the raw data (points). Asterisks indicate Benjamini-Hochberg-adjusted p-values for Mann-Whitney U tests (ns, non-significant).

To characterize these samples, we extended our previous unbiased systematic antibody profiling approach (*24*) to investigate the properties of total anti-DENV antibodies prior to severe versus non-severe disease by assessing: i) neutralization antibody titers against all four DENV serotypes and ZIKV; ii) DENV iELISA titers; iii) binding titers to DENV1-4 recombinant antigens: envelope (E), E domain III (EDIII), and non-structural protein 1 (NS1) from DENV1-4 and ZIKV); iv) Fc biophysical profiling (antibody isotypes and IgG subclasses) using recombinant antigens against DENV2, DENV3, and ZIKV; and, v) Fc effector functions (antibody-dependent complement deposition, ADCD; antibody-dependent cellular phagocytosis, ADCP; and antibody-dependent neutrophil-mediated phagocytosis, ADNP) against DENV2, DENV3, and ZIKV antigens. All the samples used in this study were from ZIKV-naïve individuals; 60 of 64 were selected from DENV epidemics that took place before the introduction of ZIKV into Nicaragua in 2016 (*12, 27*), and the 4 samples that were collected after ZIKV introduction were from children who did not have history of ZIKV infection (Table 1). Hence, we used ZIKV recombinant proteins in these assays to measure the properties of anti-DENV antibodies that cross-react with ZIKV.

In this study, we compared pre-infection samples from individuals who presented with a secondary infection that was either DF or DHF/DSS, and we analyzed our dataset in two ways: i) including all plasma samples together and against each individual antigen (DENV1, 2, 3, 4, and ZIKV) and ii) stratifying the sample set into two groups (pre-DENV2 or pre-DENV3, but not pre-DENV1 as this group only contained n=6 samples) and analyzing each group against each individual antigen (DENV1, 2, 3, 4, and ZIKV).

We first tested our samples for iELISA binding and nAb titers (Fig. 1, B to G, and fig. S1 to S2. Table 1). The iELISA assay was performed with a mixture of antigens from all four DENV serotypes (*5*). We found that iELISA binding antibody titers were not significantly associated with protection but had a p-value between 0.05 and 0.1 (p=0.0729; Fig. 1B). Antibody neutralization assays were performed to detect nAb titers of total anti-DENV antibodies (Fig. 1, C to F), as opposed to only type-specific or cross-reactive antibodies, due to sample volume limitations. nAb titers were detected but were not correlated with protection when using mature DENV1, DENV2, DENV3, and DENV4 virions. As expected (*28, 29*), this late convalescent dengue sample set did not neutralize ZIKV virions (Fig. 1G). We also did not observe statistical significance for either iELISA binding or nAb titers when we stratified the samples into pre-DENV2 and pre-DENV3 infection groups (fig. S1 and S2) In agreement with our previous study (*24*), these data suggest that, in smaller sample sets, iELISA and neutralization assays are highly variable and may not always predict protection.

### Fc biophysical features and effector functions correlate with protection from severe dengue disease

Classical correlates of protection for DENV infection and disease mostly measure the ability of the Fab portion of the antibody to bind and neutralize infectious viral particles. It is less known whether the Fc regions of anti-DENV antibodies possess protective roles against severe dengue disease and whether they may serve as additional candidates for biomarkers of protection. To investigate this, we first performed biophysical profiling of the Fc characteristics of anti-DENV antibodies by assessing different antibody isotypes (total IgA, IgM, and total IgG) and IgG subclasses (IgG1, IgG2, IgG3, and IgG4). We evaluated their binding to recombinant E, EDIII, and NS1 proteins of DENV1-4 and ZIKV, which were conjugated to microspheres used in a Luminex-based assay. We characterized the antibody binding profile by fitting a dose-response curve to estimate the area under the curve (AUC) of binding to each tested antigen and antibody isotype/subclass by participant. We then conducted logistic regression analyses to determine the correlation between the pre-existing antibody profile and the subsequent development of DHF/DSS. Each antibody and Fc effector characteristic was analyzed using a separate regression model. Within each model, adjustments were made for the incoming serotype and the DENV infection history of the individuals, applying false discovery rate (FDR) to correct for multiple hypothesis testing (Fig. S3).

Comparing the AUC between children who developed DF or DHF/DSS, we found that total IgG, IgG1, IgG2, IgG3, IgG4, and IgA, but not IgM correlated with protection from severe dengue disease (Fig. S3A). We analyzed the entire sample set against each DENV serotype and ZIKV antigen, as well as pre-DENV2 and pre-DENV3 infection sample sets against the respective incoming serotype. Total IgG and IgG1 antibodies were correlated with protection only when anti-NS1 antibodies were analyzed. IgG2, IgG3, IgG4, and IgA antibodies correlated with protection with most DENV and ZIKV antigens, as well as in the incoming serotype analysis. Of note, three IgG subclasses (IgG2, IgG3, and IgG4) were apparently more highly correlated with protection than total IgG and IgG1 (Fig. S3A), raising the possibility of antigen-antibody prozone effect in which an excess of antibody amount may mask statistically significant results. Finally, although this DENV sample set is from ZIKV-naïve individuals, most antibodies cross-reacting to ZIKV antigens were correlated with lower risk of severe disease. These results demonstrate that DENV infection generates cross-reactive antibodies against E, EDIII, and NS1 that correlate with protection from severe disease.

Next, we assessed whether the Fc effector functions ADCD, ADCP, and ADNP associate with protection (Fig. S3B, and fig. S4 to S7). We performed the functional assays using fluorescent beads conjugated to recombinant E and NS1 proteins from DENV2, DENV3, and ZIKV. We included ZIKV antigen in the functional assays since we had observed that cross-reactive antibodies binding to ZIKV antigen correlated with protection in the biophysical profiling (Fig. S3A). We conducted a bivariate analysis and multivariate logistic regression analyses adjusting by infection history and incoming serotype to determine the association between the pre-existing Fc effector functions and development of DHF/DSS (Fig S3 B and fig. S4). We found that all three effector functions tested were associated with protection (Fig. S3B). We analyzed the three pre-DENV1, -DENV2, and -DENV3 sample sets together and found that ADCD was mostly correlated with protection when cross-reacting anti-ZIKV E and NS1 antibodies were assessed (Fig. S3B). ADCP was correlated with protection when using DENV2 and DENV3 E and NS1 antigens, while ADNP was only correlated with protection with incoming serotype and E antigen (Fig. S3B).

Stratification by analyzing pre-DENV2 and pre-DENV3 infection groups individually showed that ADCD associated with protection in the pre-DENV3 sample group when using DENV3 E antigen (fig. S5). In contrast, there was no association of ADCD with protection in the pre-DENV2 group with either DENV or ZIKV antigens (fig. S5). On the other hand, ADCP was associated with protection in both pre-DENV2 and pre-DENV3 sample groups, depending on the antigen, and with anti-ZIKV E and NS1 antibodies in the pre-DENV2 sample group (fig. S6). ADNP was associated with protection in the pre-DENV2 sample group (fig. S7). ZIKV antigens were not tested in the ADNP assay. Altogether, these analyses suggest that Fc effector polyfunctionality may contribute to prevent symptomatic DENV infections evolving from DF to more severe DHF/DSS. Although the sample size is small in the stratified analyses and, therefore, interpretations should be taken with caution, these data suggest that Fc effector functions may be related with protection depending on the history of DENV infections as well as the serotype of incoming infections. In addition, our results indicate that a subset of anti-DENV antibodies cross-reacting to ZIKV antigens have Fc effector functions that associate with protection from DHF/DSS.

### Complement-mediated virolysis with ZIKV-cross-reactive antibodies strongly correlates with protection

Previously, we and others have used an antibody-dependent complement-mediated virolysis assay that successfully correlated protection from human immunodeficiency virus 1 (HIV-1) and from symptomatic DENV3 secondary infections (*24, 30*). Here, we performed a similar assay using virions from DENV, ZIKV, and YFV to verify whether the ability of antibodies to mediate virolysis was correlated with protection in this study (Fig. 2, fig. S8). By using two plasma dilutions (1:10 and 1:30), we found that all pre-infection samples induced virolysis of mature DENV2 and DENV3 virions (Fig. 2, A and B). However, we observed no statistically significant differences when comparing individuals who subsequently developed DF versus DHF/DSS. We then stratified the samples between those that were pre-secondary DENV2 and pre-secondary DENV3 (fig. S8, A to H). In support of our previous manuscript (*24*), we found that the association with protection of pre-secondary DENV3 samples incubated with DENV3 virions in the virolysis assay trended toward significance (p=0.087; dilution 1:30; fig. S8D), but pre-secondary DENV2 samples incubated with either DENV2 or DENV3 virions were not associated with protection.

**Fig. 2.**
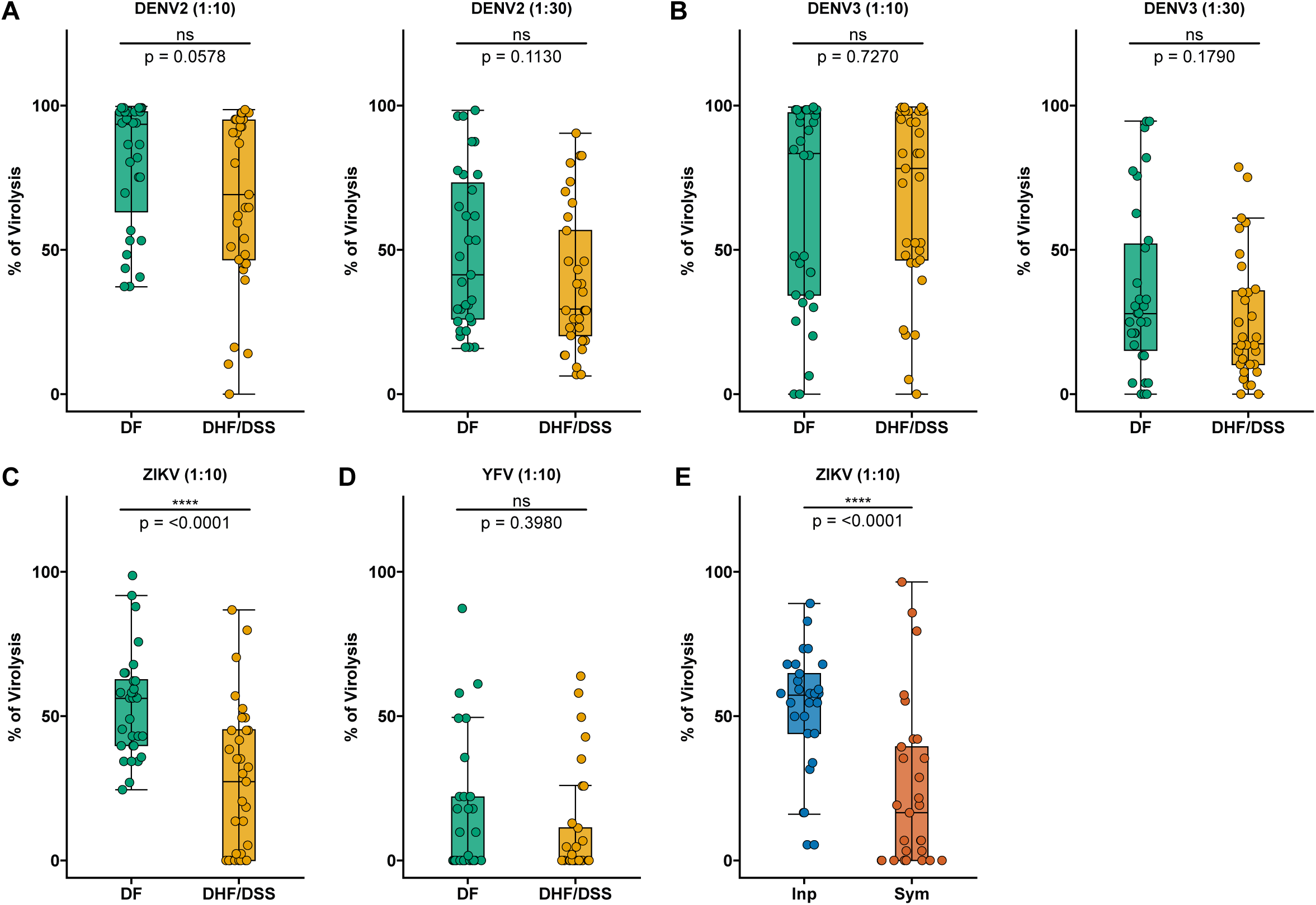
Complement-mediated virolysis with ZIKV-cross-reactive antibodies is associated with protection from subsequent DHF/DSS and symptomatic DENV infection. (A-D) Pre-infection plasma samples prior to DF (green; n=31) vs. DHF/DSS (gold; n=33) were incubated with mature DENV2, mature DENV3, ZIKV, and YFV virions. (A,B) Left panel, 1:10 dlution of plasma; right panel, 1:30 dilution of plasma. (E) Pre-inapparent (blue circles; n=30) and pre-symptomatic (orange circles; n=29) secondary DENV3 infection plasma samples were incubated with ZIKV virions. After incubation with plasma samples in the presence of guinea pig complement, mixtures were freeze-thawed once, followed by RNAse A digestion and RNA extraction from intact virions. Virion lysis was quantified by measuring viral RNA via RT-qPCR (to measure the number of genome copies) and calculated as percentage (%) of virion lysis as described in Methods. Shown are median NT_50_ (middle line), 25^th^ to 75^th^ percentile (box), and 5^th^ to 95^th^ percentile (whiskers) as well as the raw data (points). Asterisks indicate Benjamini-Hochberg-adjusted p-values for Mann-Whitney U tests (****p < 0.0001, and ns, non-significant).

As our biophysical profiling and Fc effector function assays showed that anti-DENV antibodies from ZIKV-naïve individuals that cross-reacted to ZIKV antigens were correlated with protection from DHF/DSS (Fig. 1 and S3), we performed a virolysis assay using virions from ZIKV and the more distantly related YFV (Fig. 2, C and D, and fig. S8, E to H). Importantly, there is no circulation of YFV nor widespread YFV vaccination in Nicaragua (A. Balmaseda, personal communication). Hence, all our samples were naïve to both ZIKV and YFV, allowing us to explore the properties of anti-DENV antibodies that cross-react with other flaviviruses. Although these DENV late convalescent samples did not neutralize ZIKV virions (Fig. 1G), we found that they induced virolysis of ZIKV and YFV virions but, as expected, to a lesser extent than they were capable of inducing virolysis against DENV virions (Fig. 2, C and D). Interestingly, however, ZIKV virolysis mediated by anti-DENV antibodies was significantly higher in children who subsequently developed DF compared to DHF/DSS (p<0.0001) (Fig. 2C), while there was no significant difference in virolysis of YFV virions (Fig. 2D). Furthermore, we stratified the sample set and found that virolysis was consistently higher in children who developed DF when using ZIKV virions, but not YFV, in both pre-secondary DENV2 (p=0.0207) and pre-secondary DENV3 (p=0.0089) samples (fig. S8, E to H).

To investigate whether the findings above would be reproduced in an independent sample set and context, we used the sample set from our previous study (*24*) collected from individuals before they experienced a secondary DENV3 heterotypic infection that was either inapparent or symptomatic. This sample set is also from individuals who were ZIKV-naïve, and we used them to perform a virolysis assay using ZIKV virions (Fig. 2E). Similar to our finding above, we found that virolysis of ZIKV virions was significantly higher (p<0.0001) in children with inapparent infections compared with symptomatic DENV3 infections. In sum, we identified that a subset of anti-DENV cross-reactive antibodies that are incapable of ZIKV neutralization but can bind and mediate and complement-mediated lysis of ZIKV virions are strongly associated with protection from secondary heterotypic symptomatic DENV3 infections, as well as from progression to DHF/DSS in secondary DENV2 and DENV3 infections.

### Virolysis is the main antibody feature correlated with protection from severe symptoms and disease

We evaluated the performance of the antibody biophysical profile and the elicited Fc effector functions in pre-infection samples to discriminate between children who subsequently developed DF versus DHF/DSS. We employed a multivariate regression model with regularization, incorporating variables that showed significant differences in the bivariate analysis, to down-select discriminant features between both clinical groups. Initially, when evaluating the classification performance of both the antibody profile and elicited Fc effector functions, regardless of the incoming serotype, our model yielded an AUC of 0.74 (95% CI 0.56-0.92) and identified ZIKV virolysis as the most relevant classification variable (Fig. 3, A and B). However, since DHF/DSS was caused by both secondary DENV3 (n=33) and DENV2 (n=25) infections, we stratified the analysis based on the incoming serotype (Fig. 3, C to F). This analysis improved the performance of the model and revealed specific differences in Fc effector functions enriched in the DF group by serotype. In the model for incoming DENV2 (AUC= 0.95, 95% CI 0.82-1.00), ADNP elicited by DENV2 E protein emerged as the most relevant feature, alongside ADCP against ZIKV E and virolysis of ZIKV virions (Fig. 3, C and D). In the model for incoming DENV3 (AUC = 0.88, 95% CI 0.71-1.05), ZIKV virolysis and binding of IgG2 antibodies to DENV3 E protein were the most relevant protective variables, enriched in the DF group (Fig. 3, E and F).

**Fig. 3.**
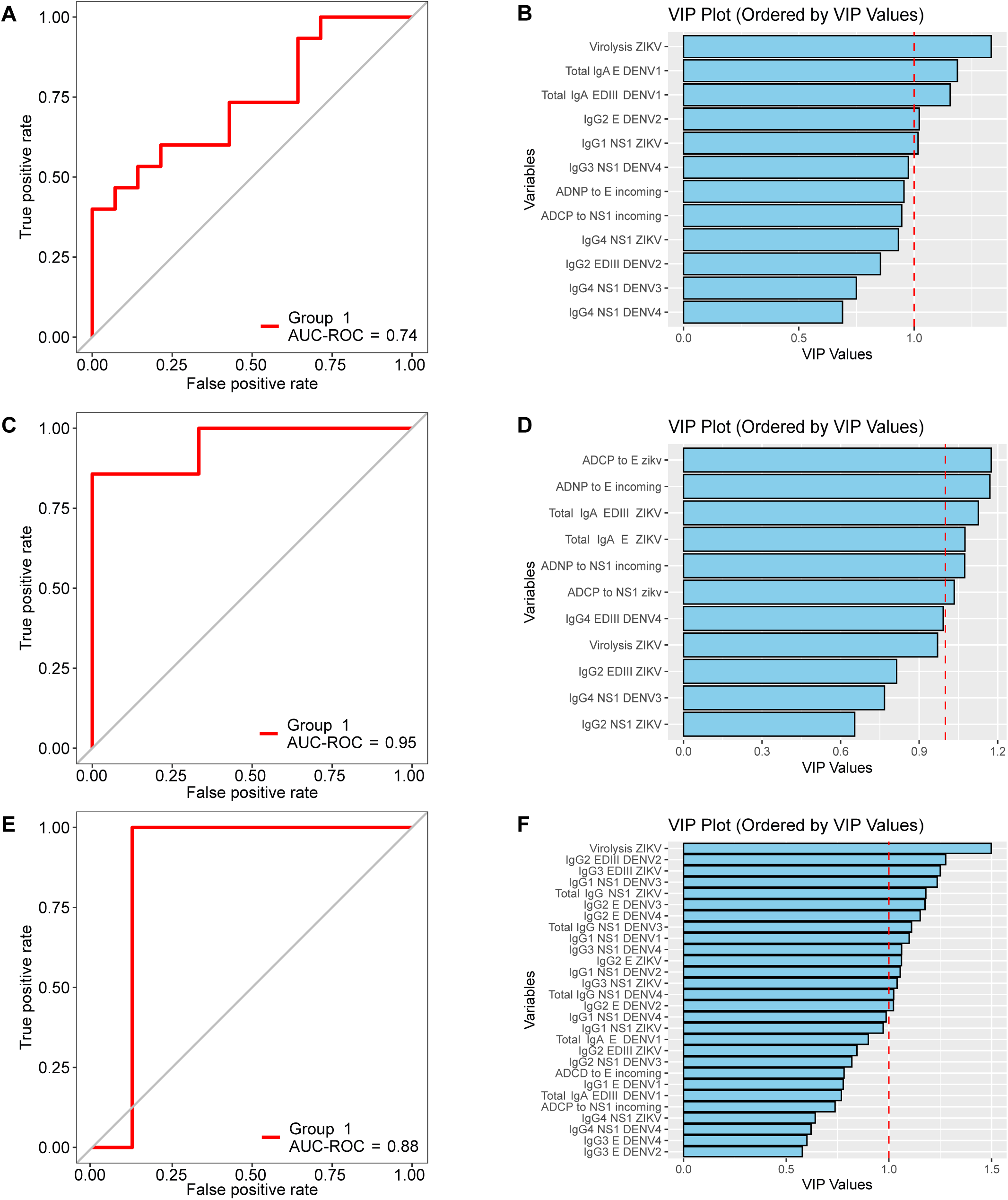
Variables discriminating DHF/DSS and DF vary depending on incoming serotype but overall, ZIKV virolysis is the main discriminating antibody feature. Multivariate regression models with regularization, incorporating variables that showed significant differences in the bivariate analysis, were used to identify discriminant features between subsequent DF vs. DHF/DSS. (A-B) Multivariate regression model with regularization without stratifying by incoming serotype. (C-D) Multivariate regression model with regularization stratified for incoming DENV2. (E-F) Multivariate regression model with regularization stratified for incoming DENV3. (A,C,E) Receiver operator curve (ROC) analysis of the regression models, with the area under the curve (AUC) values displayed. (B,D,F) Variable of importance plot (VIP) derived from the regression models. The red dashed line indicates a VIP value of 1. The regression model without stratification (regardless of the incoming serotype) identified ZIKV virolysis as the most relevant variable enriched in DF (A-B). The analysis stratified by incoming serotype of pre-DENV2 (C-D) and pre-DENV3 (E-F) infection improved the performance of the model and revealed differences in variables discriminating subsequent DHF/DSS vs. DF, including ZIKV virolysis.

Finally, we investigated which of these Fc effector functions correlated with protection from developing severe symptoms and signs associated with dengue disease (Fig. 4). After correction by FDR, only ZIKV virolysis correlated with lower odds of developing not only DHF/DSS but also relevant clinical signs, such as thrombocytopenia, hemorrhagic manifestations, and plasma leakage even after adjusting for incoming serotype, infection history, and time since last infection (Fig. 4). Our analyses underscore the potential of various antibody Fc properties present in pre-infection plasma antibodies to identify individuals at lower risk of progressing to DHF/DSS and developing severe symptoms and signs. In particular, virolysis mediated by a subgroup of ZIKV cross-reactive antibodies appear to be one of the most important antibody features correlated with protection from severe disease and clinical manifestations.

**Fig 4.**
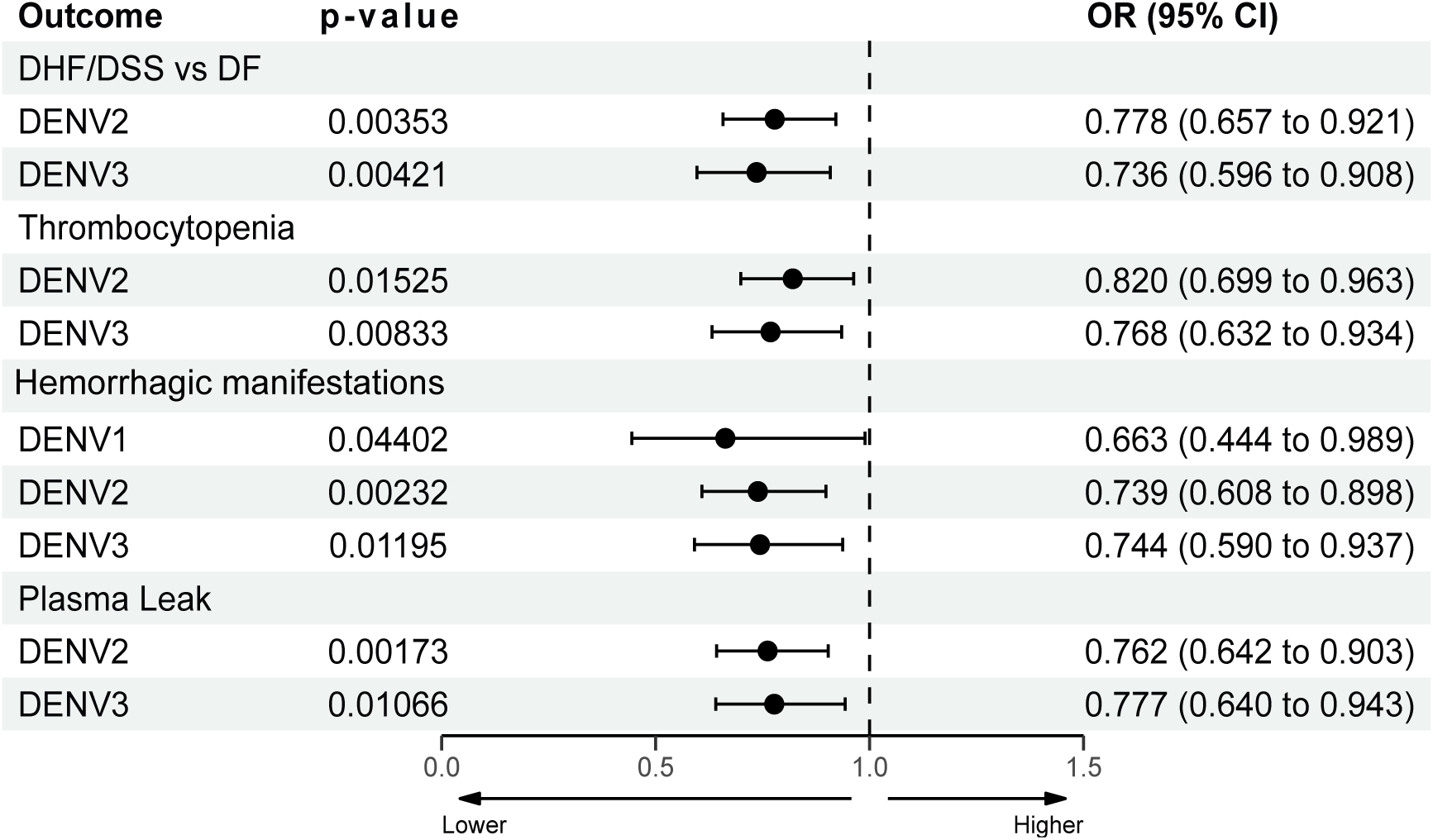
Complement-dependent virolysis mediated by ZIKV-cross-reactive antibodies is correlated with protection from symptoms and signs of DHF/DSS. Forest plot showing the adjusted odds ratios (OR), confidence intervals (CI), and p-value of the correlation between DHF/DSS or symptoms of DHF/DSS (thrombocytopenia, hemorrhagic manifestations, and plasma leak) and ZIKV virolysis. Odds ratios (OR) were adjusted by incoming serotype (interacting variable), infection history, and time since last infection. OR are shown with 95% confidence intervals (95% CI).

## DISCUSSION

There is an urgent need for the development of tools that enable identification of individuals with prior DENV infection exposed to DENV who are at risk of developing severe disease upon a secondary DENV infection. Here, we evaluated the binding profile, nAb titers, and Fc effector functions elicited by antibodies in plasma samples obtained from children with history of DENV infection(s) but before a subsequent secondary heterotypic symptomatic infection (DF versus DHF/DSS) and their potential to identify children at lower risk of developing severe disease. We show that complement-mediated ZIKV virolysis of plasma obtained prior to a secondary DENV infection correlated with protection from subsequent DHF/DSS. Importantly, this correlation of pre-infection ZIKV virolysis remained after adjusting by infection history, time since last infection, and incoming infecting serotype (DENV1, DENV2, and DENV3), and applied to DHF/DSS as well as signs and symptoms of severe disease (thrombocytopenia, hemorrhagic manifestations, and plasma leakage).

Currently, neutralization assays have been the main method used to assess protection in vaccine trials and natural infection studies, with the caveat that nAb titers do not always correlate with protection from disease in humans. Multiple parameters (e.g., virion maturation state, cell line substrate, infection history, and incoming infecting serotypes) have been shown to affect neutralization assays, highlighting the need to control assay conditions carefully and to develop novel methods to assess protection (*22, 36*). The complex nature of anti-DENV antibodies and the lack of tools to indicate individuals protected from disease have led to major shortcomings in vaccine development. It has been well documented that DENV infections generate cross-reactive antibodies that can protect or enhance subsequent heterotypic DENV infections and/or infections of closely antigenically related flaviviruses (*5–7, 12–15, 20*). Various factors contribute to these contrasting characteristics, including immune serostatus (e.g., past infections and/or vaccinations), time post-infection, order of infection, serotypes, viral strains, antibody post-translational modifications, and host genetics (*6, 11, 15, 22, 29, 33–35*). A novel biomarker that allows identification of individuals at risk of severe disease, independent of the infecting serotype, the infection history, or the time since last infection, may have the potential to facilitate evaluation of vaccine safety and efficacy. Here, we show that cross-reactive antibodies against the related ZIKV virion, produced after DENV infection in ZIKV-naïve children, that induced ZIKV virolysis were correlated with protection against severe disease across serotypes.

To investigate novel candidate antibody markers that could potentially be measured alongside binding and nAb titers, in our previous (*24*) and present studies, we selected pre-infection samples from our cohort in Nicaragua prior to a subsequent secondary DENV infection resulting in symptomatic versus inapparent infection or severe versus non-severe disease, respectively. In these sample sets, we observed that nAb titers did not correlate with protection. We assessed other antibody properties, such as Fc biophysical features and effector functions that associated with protection. In both studies, we identified multiple similar anti-DENV antibody isotypes, subclasses, and Fc-effector functions that correlated with protection, generating evidence in a pediatric cohort that the Fc regions of anti-DENV antibodies contain properties that can protect children during secondary DENV infections. Other studies have reported Fc afucosylation associated with severity in post-infection samples collected during the acute phase when inflammatory reactions are ongoing (*37*). As our analysis was performed in pre-secondary infection samples, during a resting period as opposed to an acute inflammatory state, this may have enabled identification of protective properties of the Fc regions of anti-DENV antibodies. Our data on IgG subclasses and polyfunctionality suggest the possibility that different subclasses may work in cooperation to mediate protection and that activating and inactivating features of the Fc regions could be important for triggering a strong antiviral immune response during early stages of viral infection while preventing an undesired exacerbated response that could lead to severe symptoms. This concept has been observed, for example, in a dengue study in Cambodia revealing that asymptomatic individuals presented with strong activation of adaptive responses while preventing its overactivation through concomitant activation of feedback mechanisms of control (*32*).

Another consistent finding between our previous and current studies was the observation that, despite the overall low levels of circulating IgG4 antibodies, protected individuals had significantly higher titers of IgG4 antibodies. Interestingly, two other recent studies also identified high levels of IgG4 antibodies upon multiple doses of COVID-19 and HIV-1 vaccination regimens and an association with reduced Fc-effector functions (*33, 34*). These studies show that IgG4 levels increase in parallel to IgG1 responses, potentially representing a subpopulation of antibodies with more neutralizing capacity and, perhaps, class-switched to restrain potential pathology. However, the parallel augmented IgG1 response may play a dominant role in driving viral protection via accelerated pathogen clearance and destruction. These observations in natural infections and vaccine studies may suggest that during DENV infections, repeated exposures in endemic regions and may lead to increased levels of circulating IgG4 in protected individuals. Given the low volumes of our pediatric cohort samples, we were unable to directly test here the putative protective roles of IgG4 in DENV infections, but possible mechanisms could be associated with increased neutralization potential; contribution to a balanced Fc-effector function response that may prevent overactivation of the immune system; competition for binding to epitopes associated with ADE; and potential roles associated with the bi-specific nature of IgG4, among others (*35*). Future studies are needed to clarify which factors can increase the titers of circulating IgG4 antibodies in infectious diseases and whether these are a surrogate of enhanced antibody function or mechanistically linked to protection.

Antibodies binding to the three antigens tested (E, EDIII, and NS1) were associated with protection, including anti-E and -NS1 antibodies possessing Fc effector functions (ADCD, ADCP, and ADNP). As in our previous study using only pre-secondary DENV3 samples (*24*), we also found ADCD to be associated with protection. In the present study, which includes pre-secondary DENV1, DENV2, and DENV3 infection samples, after performing a stratification analysis by DENV serotype, we found that ADCD was correlated with protection especially in the pre-secondary DENV3 infection sample group, using E and NS1 recombinant antigens. In contrast, we found that nAb titers are more correlated with protection in pre-DENV2 compared to pre-DENV3 infection samples (*36*), raising the possibility that protection against different DENV serotypes may be associated with different antibody functions. When the ADCD assay was performed with ZIKV E and NS1 antigens, protection was observed when all samples were analyzed together, but not when the analysis was stratified by pre-DENV2 and pre-DENV3 subgroups. However, when we used a virolysis assay to substantiate the ADCD assay findings, we observed that virolysis of ZIKV virions was strongly correlated with protection when all samples were analyzed together, as well as when pre-DENV2 and pre-DENV3 sample sets were stratified and analyzed separately. It is possible that the virolysis assay conducted with ZIKV virions indicates the existence of protective antigenic sites present (or enriched) on the surface of virions that are lacking on recombinant antigens such as the ones used in the ADCD assay. Our analyses revealed that the virolysis assay with ZIKV virions was one of the main antibody properties correlating with protection from DHF/DSS, while being the only feature correlated with protection from specific severe clinical manifestations. Thus, the virolysis assay conducted with ZIKV virions appears to have the potential to identify individuals at lower risk of severe disease upon a secondary DENV infection across different serotypes and infection histories.

Our dengue sample set is derived from ZIKV-naïve individuals; thus, it is interesting to note that this subset of anti-DENV antibodies was incapable of neutralizing ZIKV but was able to bind and mediate complement-dependent lysis of ZIKV virions. ZIKV virolysis was correlated with the outcome of future secondary heterotypic DENV infections in two independent sample sets, while virolysis with DENV virions provided none or only a weaker association with protection. It is possible that the complexity associated with the contrasting properties of various anti-DENV antibodies in an *in vitro* assay with DENV virions may mask the protective phenotype associated with a small population of protective ZIKV-cross-reacting antibodies. In humans, other immune responses may complement and augment the protective role of this subset of complement-fixing antibodies. Whether complement lysis alone is mechanistically linked to viral clearance, or whether complement deposition on the surface of the virion may accelerate viral uptake and rapid lysosomal destruction remains unclear but may provide insights regarding protective mechanisms prior to secondary DENV infections. Furthermore, as for neutralization assays showing improved capacity to predict protection when type-specific antibodies are measured (*19, 22, 23*), the virolysis assay may also be improved when a subset of anti-DENV antibodies (e.g., ZIKV cross-reacting antibodies) is measured. Unlike type-specific neutralization assays, depletion of antibodies prior to the virolysis assay was not required, and small volumes of plasma samples were used, highlighting another advantage of this technique. Of note, YFV 17D-cross-reacting antibodies were not associated with protection, suggesting that ZIKV-but not flavivirus-cross-reacting antibodies are likely to be responsible for the phenotype noticed in this study. Future studies are needed to explore the antigenic sites associated with protection and identify antigen-specific antibodies circulating in the plasma via de novo MS/MS sequencing (*37, 38*).

Antibodies targeting the Envelope Dimer Epitope (EDE) were shown to facilitate broad neutralization across flaviviruses (*39, 40*). EDE antibodies target a quaternary epitope formed through the dimerization of E proteins, crucial for viral entry into host cells (*41, 42*). It would be interesting to investigate the potential of these anti-EDE antibodies to promote virolysis, which might also contribute to their protective effects, clarifying their functional mechanisms. In addition, independent studies revealed a broadly DENV-ZIKV neutralizing antibody clone that was recently engineered to remove binding to FcyRs – in order to prevent the possibility of ADE – while still allowing complement activation (*43, 44*). The latter study demonstrated that complement activation mediated by this engineered antibody had protective properties in a DENV2 mouse model. Unlike the antibodies described in all these previous studies, the ZIKV-cross-reacting antibodies identified here were unable to cross-neutralize ZIKV virions, suggesting that different (or even novel) epitopes may be associated with complement-mediated protection. Further experiments are necessary to identify the epitope(s) targeted by these antibodies and to validate their ability to induce virolysis. This continued investigation will enrich our understanding of the antibody repertoire generated upon DENV infection and of the complex interplay of antibody-mediated protection and pathogenesis in flavivirus infections, potentially guiding the development of more effective vaccines and therapeutic interventions.

One potential shortcoming of our virolysis assay is that during the ZIKV epidemic, many individuals were exposed to ZIKV, and the presence of actual anti-ZIKV antibodies (as opposed to anti-DENV antibodies that cross-react with ZIKV) in the population may affect the assay results and interpretation. Currently, we believe this assay may have better potential in a population not affected by ZIKV, and we will assess this in future studies. We are currently evaluating the virolysis assay in dengue cohorts of natural infection from other geographical regions and in samples from vaccine trials. Given the limitations and complexity in developing other bioassays, we did not assess further in this study the other protective Fc functions (e.g., ADCP and ADNP). In a separate note, we and others previously shown epidemiological evidence that past DENV infections protect from future ZIKV disease (*12, 13*). Considering our findings that some anti-DENV antibodies are poorly neutralizing, but lead to virolysis of ZIKV virions, we also consider the possibility that virolysis may be a potential mechanism of protection associated with this epidemiological observation.

In sum, we identified anti-DENV antibodies that cross-react with ZIKV virions and were strongly correlated with protection from symptomatic infection and severe disease due to secondary heterotypic DENV2 and DENV3 infections. This protection associated with the ability of ZIKV-cross-reactive antibodies to induce lysis of ZIKV virions, mediated by the complement pathway. The virolysis assay has emerged as a potential biomarker of protection in natural infection studies and possibly vaccine trials. The future identification and characterization of the antigenic sites and sequencing of these protective antibodies could contribute to the rational design of next-generation vaccine candidates and therapeutics.

## MATERIALS AND METHODS

### Ethics statement

The human subjects protocol for the Pediatric Dengue Cohort Study (PDCS) was reviewed and approved by the Institutional Review Boards (IRB) of the University of California, Berkeley (2010-09-2245), the University of Michigan (HUM00091606), and the Nicaraguan Ministry of Health (CIRE-09/03/07-008). Parents or legal guardians of all participants provided written informed consent, and participants 6 years of age and older provided assent. All participants live in District 2 of Managua, Nicaragua, the catchment area for the Health Center Socrates Flores Vivas (HCSFV).

### Samples and study population

The PDCS is an ongoing open prospective cohort of ∼4,000 children 2-17 years old that was initiated in 2004 in Managua (*26*). Blood samples are collected from healthy participants annually, and additional blood samples are collected from symptomatic suspected dengue cases at the acute (0-6 days post-onset of symptoms) and convalescent (14-21 days post-onset of symptoms) phase. DENV infections are evaluated and recorded yearly by comparing healthy annual plasma samples collected in two consecutive years side-by-side by inhibition enzyme-linked immunosorbent assay (iELISA) (*5*). A ≥ 4-fold increase in iELISA titer between annual samples is considered as indication of an infection (REF).

Symptomatic dengue cases were confirmed by detection of DENV RNA by RT-PCR, real-time RT-PCR and/or virus isolation in the acute-phase sample; seroconversion of DENV-specific immunoglobulin M antibodies in paired acute- and convalescent-phase samples; or seroconversion or a ≥ 4-fold increase in antibody titer by iELISA between acute and convalescent sera (*5, 26, 45*). The serotype of symptomatic DENV infections was determined by RT-PCR or real-time RT-PCR (*46, 47*).

To investigate biomarkers of protection associated with a secondary heterotypic DENV infection, we selected 64 children who had previously experienced DENV infection and who subsequently developed a symptomatic infection (n = 31) or severe infection (n = 33) and analyzed their pre-infection plasma samples collected the year before the DF or DHF/DSS episode. The selection involved identifying all individuals from the PDCS between 2004 and 2019 who had a DHF/DSS episode and a healthy sample taken within 12 months before their secondary infection. The DHF/DSS group included 7 cases of DENV1, 25 of DENV2, and 23 of DENV3. DF controls were randomly matched with severe cases by serotype and infection history, applying frequency matching. Cases and controls were restricted to individuals without prior exposure to the incoming serotype or ZIKV as confirmed by various methods such as reverse transcriptase-polymerase chain reaction (RT-PCR), nAb titers, epidemiological data, inhibition enzyme-linked immunosorbent assay (iELISA) binding antibody titers, ZIKV BOB ELISA, and binding to envelope (E) protein domain III (EDIII) in a Luminex assay (*5, 14, 36, 48*). Samples obtained after a single DENV infection are referred to as post-primary infection samples, while samples obtained after two or more prior DENV infections are referred to as post-secondary infection samples (Fig.1). No significant differences in age, sex, time since last infection, pre-existing DENV antibody levels, or infection year were found between the DHF/DSS cases and DF controls (Table 1).

Samples in this study are from pediatric participants in our long-term cohort study and are therefore very limited in volume. Thus, these materials are subject to restriction based on limited availability and require approval from the IRB, Dr. Harris and Nicaraguan researchers. For more information, please contact Dr. Eva Harris and/or the committee for the Protection of Human Subjects at University of California Berkeley (510-642-7461).

### Cell lines

Vero cells (ATCC CCL-81), Vero cells stably expressing human furin (Vero-Furin; kind gift of Drs. Victor Tse and Ralph Baric, University of North Carolina (UNC), Chapel Hill)(REF), were maintained in minimum essential medium (MEM; Gibco) with 5% fetal bovine serum (FBS) supplemented with 1X non-essential amino acids (Gibco), 1X sodium pyruvate (Gibco), and 1X penicillin-streptomycin (Gibco) at 37°C in 5% CO_2_.

### Virus production

DENV1 (5575.10a1SPD1), DENV2 (8891.12a1SPD2), DENV3 (6629.10a1SPD3), DENV4 Sri Lanka 92 (KJ160504.1) and ZIKV (Nica 2-16) are clinical isolates from Nicaragua, isolated on C6/36 cells and passaged twice in Vero cells prior to production of working stocks.). Mature viruses were grown in Vero-furin cells in the same growth medium described above. Virions were harvested 5 days post-infection and frozen at 80°C until use.

### Neutralization Assay

The focus reduction neutralization test (FRNT) assay was conducted using Vero cells. Briefly, cells were seeded in a 96-well plate one day before infection. Plasma samples were serially diluted and mixed with an equal inoculum of DENV1-4 or ZIKV (equivalent to 40–50 FFU/well) at a volume ratio of 1:1 to form immune complexes. The virus-antibody mixture was incubated for 1 hour (h) at 37°C in 5% CO_2_. Cell growth medium was removed, and the virus-antibody mixture was added to Vero cells and incubated for 1 h at 37°C in 5% CO_2_, after which the plates were incubated for 30 h. Foci were developed using True Blue HRP substrate (KPL) and were counted using an CTL Immunospot analyzer (Cellular Technology, Ltd.). Foci counts were fitted using a variable slope dose-response curve using Graphpad, and neutralizing antibody titer (NT_50_) values were calculated with constrained top and bottom values of 100 and 0, respectively. Percent relative infection was calculated as a ratio of foci counts in each serial dilution to the foci counts from the final serial dilution of each sample. All samples were run in duplicate, with reported values required to have an R^2^ > 0.85 and a Hill slope > 0.5. Neutralization titers were measured against the heterotypic incoming serotype of each participant.

### Viral antigens

Recombinant E (recE) and NS1 proteins from DENV1-4 and ZIKV were commercially acquired from Native Antigen Co. (United Kingdom) and used for biophysical profiling and Fc effector function assays. The recombinant domain III of DENV1-4 and ZIKV E protein (EDIII) were site-specifically biotinylated and provided as a gift by Drs. Aravinda de Silva and Lakshmanane Premkumar (UNC, Chapel Hill).

### Antibody subclass and isotype binding

Antibody subclass and isotype binding was determined using a multiplex Luminex assay, as previously described (*24*). Briefly, recE and NS1 from DENV1-4 and ZIKV were random biotinylated according to the manufacturer’s instructions (EZ-Link™ Sulfo-NHS-LC-LC-Biotin, No-Weigh™ Format; Thermo Fisher Scientific) and desalted using Zeba Spin Columns (Thermo Fisher Scientific). The random biotinylated recE, NS1, and the site-specific biotinylated EDIII from DENV1-4 and ZIKV were conjugated to avidin-coated MagPlex Luminex beads. Immune complexes were formed in 384-well plates by mixing the appropriately diluted plasma (six four-fold dilutions) with antigen-coupled microspheres for 90 minutes at 37°C, shaking at 1200 rpm. After incubation, plates were washed using an automatic magnetic washer (Tecan Hydrospeed) with phosphate-buffered saline (PBS) containing 0.1% bovine serum albumin (BSA) and 0.02% Tween 20. Antigen-specific subclass/isotype binding was detected using PE-coupled mouse anti-human detection antibodies against IgA (SouthernBiotech #2050-09), IgM (SouthernBiotech #9020-09), total IgG (SouthernBiotech #9040-09), IgG1 (Southern-Biotech #9052-09), IgG2 (SouthernBiotech #9060-09), IgG3 (Southern-Biotech #9210-09), and IgG4 (SouthernBiotech #9200-09). Fluorescence was acquired using an iQue3 (Intellicyt) machine. Antigen-specific antibody subclass/isotype binding was extracted as median fluorescence intensity (MFI). Then, a netMFI was calculated by subtracting the background binding of a DENV naïve sample from each test sample.

### Antibody Fc functional assays

ADCP, antibody-dependent neutrophil phagocytosis (ADNP) and ADCD were performed as previously described (57–60). For ADCP, ADNP, and ADCD, antigen was biotinylated and coupled to yellow-green or red (ADCD) neutravidin beads (Invitrogen). Immune complexes were formed by mixing coupled beads and appropriately diluted plasma and incubating for 2 h at 37°C. For ADCP, human monocytic THP-1 cells [American Type Culture Collection (ATCC) TIB-202] were added to immune complexes at a concentration of 1.25 × 105 cells/ml, and cells and immune complexes were incubated overnight at 37°C. For ADNP, white blood cells were isolated from fresh peripheral blood from healthy donors by performing ammonium-chloride-potassium lysis of red blood cells. The isolated white blood cells were then added to 96-well plates at a concentration of 2.5 × 10^5^ cells/ml, and neutrophils were identified using anti-CD66b PacBlue (BioLegend, 305112). For ADCD, lyophilized low-toxicity guinea pig complement (Cedarlane) diluted in gelatin veronal buffer with calcium and magnesium (GVB++, Sigma-Aldrich) was added to immune complexes and incubated for 20 minutes at 37°C. Fluorescein-conjugated goat IgG anti-guinea pig complement C3 detection antibody (MP Biomedicals #MP0855385) was used to detect complement deposition.

Fluorescence was acquired using an iQue3 machine (Sartorius). For ADCP and ADNP, a phagocytic score was calculated using the following formula: (percentage of bead-positive cells) x (geometric mean of MFI of bead-positive cells)/10,000. ADCD activity is reported as the MFI of C3 deposition. ADNP were run using fresh peripheral blood from two healthy donors. ADCP and ADCD were run in duplicate. The data presented reflect the average of the biological (ADNP) or technical (ADCP and ADCD) replicates.

### Virological assays in the presence of complement

The virion lysis assay was performed as previously described (*24, 30*). Briefly, plasma samples were diluted and used at final concentrations of 20, 4, or 1.3% and mixed with fresh guinea pig complement (resuspended in 1 ml of GVBLJLJ, Sigma-Aldrich; final concentration of 50%), DENV2 8891 strain virions, DENV3 strain 6629 virions, ZIKV strain Nica 2-16 virions or YFV 17D strain in RPMI 1640 (Gibco). The mixtures were incubated at 37°C for 3 h and frozen overnight at −20°C. Samples were then thawed, mixed with RNase A (final concentration, 0.77 mg/ml), and incubated at 37°C for 1 h. Reactions were terminated by incubating with proteinase K (final concentration, 0.71 mg/ml) at 50°C for 1 h. RNA was extracted and purified using the RNeasy Mini Kit (QIAGEN). Absolute quantitation of viral RNA was determined by real time RT quantitative PCR (RT-qPCR) using the following primer and probe sequences: DENV2 forward (ACAAGTCGAACAACCTGGTCCAT), reverse (GCCGCACCATTGGTCTTCTC) and probe (FAM-CCAGTGGAATCATGGGAAGAAGTCCCA-TAMRA; DENV3 forward (GGACTGGATACACGCACCCA), reverse (CATGTCTCTACCTTCTCGACTTGTCT), and probe (FAM-ACCTGGATGTCGGCTGAAGGAGCTTG-TAMRA); ZIKV forward (CCGCTGCCCAACACAAG), reverse (CCACTAACGTTCTTTTGCAGACAT) and probe (FAM-AGCCTACCTTGACAAGCAGTCAGACACTCAA-TAMRA; YFV forward (GCACGGATGTAACAGACTGAAGA), reverse (CCAGGCCGAACCTGTCAT) and probe (FAM-CGACTGTGTGGTCCGGCCCATC-TAMRA (REF – ATCC website). The Verso 1-step RT-qPCR kit (Thermo Fisher Scientific) was used following the manufacturer’s instructions. Standard curves were prepared using seven 10-fold dilutions of a DENV2 8891 synthetic fragment, DENV3 UNC3009 in-vitro transcribed RNA, ZIKV RNA standard (ATCC VR-3252SD) or YFV synthetic RNA standard (ATCC VR-3253SD). Reaction mixtures were run in a QuantStudio 3 thermocycler (Applied Biosystems), and the quantity values were used for analyses. Percentages of virion lysis were calculated using the average quantity of four dengue-naïve plasma samples that were used as negative controls. Plasma samples derived from several dengue-immune individuals were mixed to form one pool or two of DENV-positive plasma samples and were used as positive control for the assay.

### Statistical analysis

All statistical analyses were conducted using R (R Foundation for Statistical Computing, version 4.5.0). We fitted a dose-response curve using the netMFI across six 4-fold serial dilutions of plasma to estimate the (AUC) of the binding for each tested antigen, antibody subclass/isotype and participant. The AUC was compared between children who subsequently developed DF or DHF/DSS to identify significant associations after applying false discovery rate (FDR) correction for multiple comparisons. NT_50_ values were obtained using GraphPad software as indicated above. Normal distribution of data was tested using the Shapiro-Wilk test. Adjusted p-values and statistical significance of nAb titer magnitude and differential NT50 comparisons were computed using the Wilcoxon test with Benjamini–Hochberg correction.

To evaluate the classification performance of both the antibody profile and elicited Fc effector functions, we employed multivariate regression with regularization. Due to the complexity of the model and the high collinearity of the data, we reduced data dimensionality by incorporating variables that showed significant differences in the bivariate analysis. Next, we divided our sample set into a training set for model fitting and a testing set using a 50-50 split. To determine the optimal regularization parameters, we conducted cross-validation (n=10). Subsequently, only the best gamma and alpha values, which improved the AUC of the model, were evaluated on the testing set to assess performance and select the best model. Finally, the best model was employed to re-estimate the coefficients of the selected features across the entire dataset. Regularization was performed using glmnet R package v 4.1-8 (*49*).

We conducted a logistic regression analysis to determine the correlation between the pre-existing antibody profile and the development of DHF/DSS. Each antibody and Fc effector characteristic was analyzed using a separate regression model. Finally, the models that were significant after correcting by FDR were adjusted accounting for the incoming serotype (as interacting variable), the DENV infection history of the individuals, and the time since last infection.

## Supporting information

Dias Duarte et al Supplemental Figures

## Data Availability

All data pertinent to this study can be found within the paper or the Supplementary Materials. After securing approval from the UC Berkeley Committee for the Protection of Human Subjects, individual data for figure reproduction could be shared with external researchers. For data access arrangements, please contact EH at. Standard data and material transfer agreements govern all materials and data used in this study.

## List of Supplementary Materials

Figures S1-S8.

## Acknowledgments

We thank the study personnel at the Centro de Salud Sócrates Flores Vivas, the Nicaraguan National Virology Laboratory, the Hospital Infantil Manuel de Jesús Rivera, and the Sustainable Sciences Institute. We are particularly grateful to the study participants and their families.

## Author contributions

Conceptualization: AGDJ, ED, GA, EH

Investigation and Data Analysis: AGDJ, ED, JVZ, JACO, SB

Sample Selection: JVZ, GK, AB, EH

Visualization: JVZ, JACO

Funding acquisition: GA, EH

Project administration: AB, GA, EH

Supervision: AB, GA, EH

Writing – original draft: AGDJ, ED, JACO, EH

Writing – review & editing: All authors.

