## Supplementary material for "Complement-dependent virion lysis mediated by dengue-Zika virus cross-reactive antibodies correlates with protection from severe dengue disease": Dias Duarte et al Supplemental Figures

**This pdf file includes:**

Figures S1 to S8

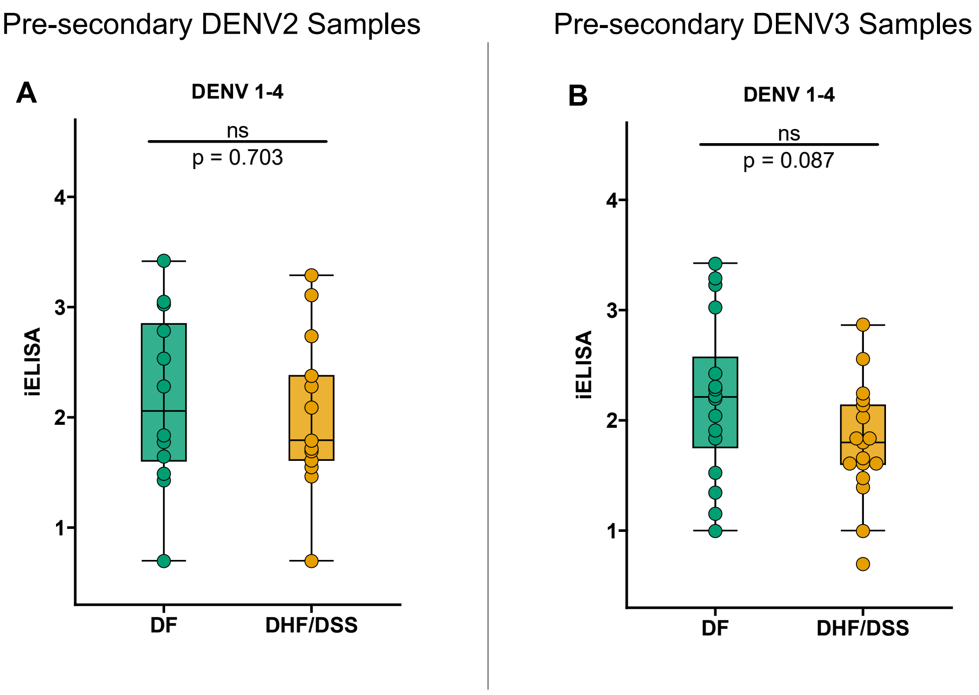


**Fig. S1. DENV iELISA titers in pre-infection samples from participants who experienced subsequent DF vs. DHF/DSS stratified by incoming serotype.** (A) Pre-secondary DENV2 samples. (B) Pre-secondary DENV3 samples. Shown are median iELISA titers (middle line), 25$^{th}$ to 75$^{th}$ percentile (box), and 5$^{th}$ to 95$^{th}$ percentile (whiskers) as well as the raw data (points). Asterisks indicate Benjamini-Hochberg-adjusted p-values for Mann-Whitney U tests (ns, non-significant).

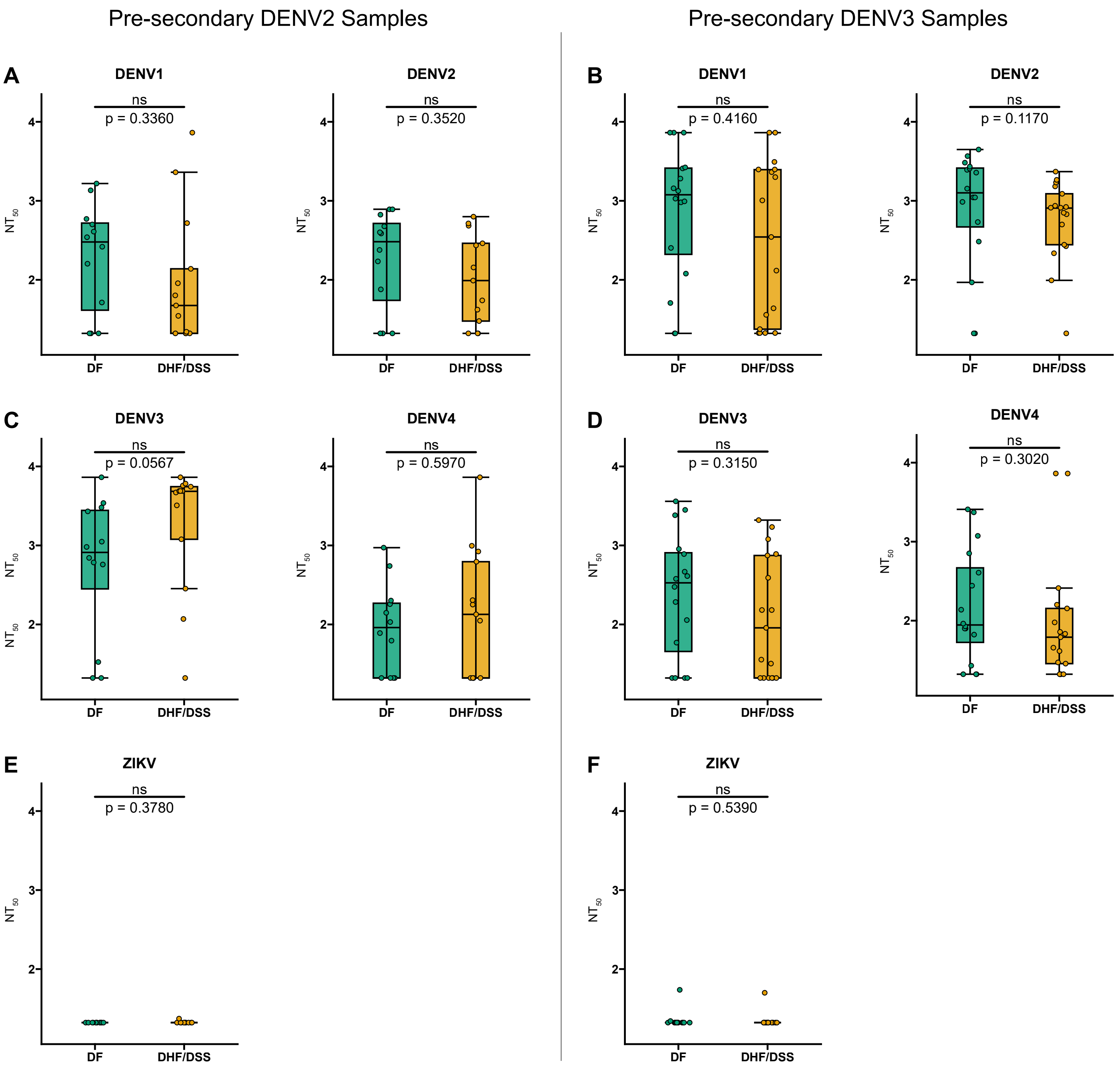


**Fig. S2. Neutralizing antibody titers in pre-infection samples from participants who experienced subsequent DF vs. DHF/DSS stratified by incoming serotype.** (A,C,E) Pre-secondary DENV2 samples. (B,D,F) Pre-secondary DENV3 samples. (A,B) DENV1, left panel; DENV2, right panel. (C,D) DENV3, left panel; DENV4, right panel. (E,F) ZIKV. Shown are median $NT_{50}$ (middle line), 25th to 75th percentile (box), and 5th to 95th percentile (whiskers) as well as the raw data (points). Asterisks indicate Benjamini-Hochberg-adjusted p-values for Mann-Whitney U tests (ns, non-significant).

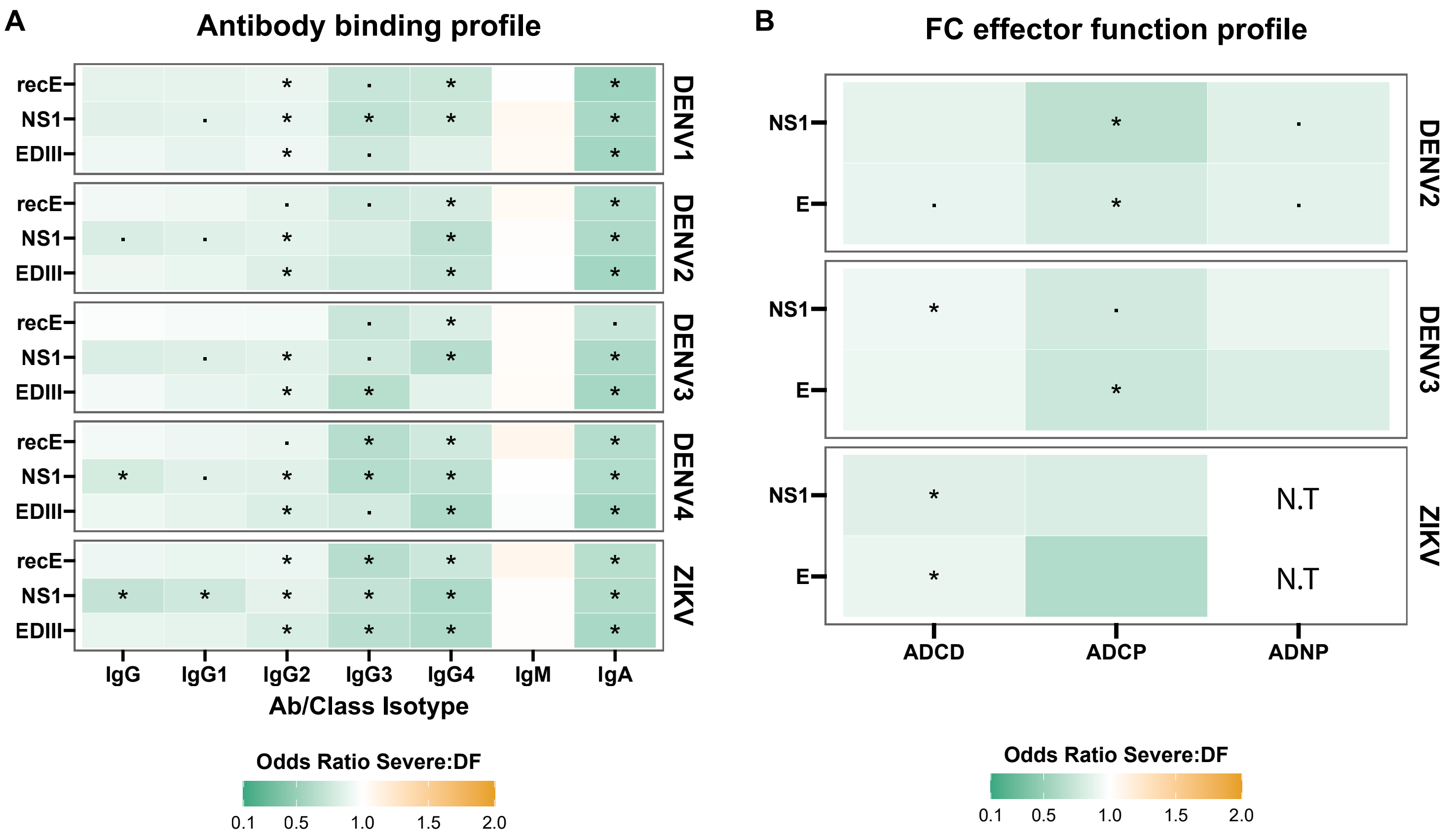


**Fig. S3. Biophysical and Fc effector functions of anti-DENV polyclonal antibodies.** (A-B) Heatmap of odds ratios demonstrating the correlation between antigen-specific antibody isotype levels and Fc effector functions for antibodies targeting the envelope (E), E domain III (EDIII), and nonstructural protein 1 (NS1) of DENV1-4 and ZIKV, and the likelihood of developing Dengue Hemorrhagic Fever (DHF) or Dengue Shock Syndrome (DSS) upon subsequent infection. (A) Antibody isotypes were measured by a Luminex-based assay and are shown as median fluorescence intensity (MFI). (B) Cellular Fc effector functions investigated were antibody-dependent cellular phagocytosis (ADCP) and antibody-dependent neutrophil phagocytosis (ADNP). Heatmaps of odds ratios were derived from multivariate analysis accounting for infection history for biophysical (A) and Fc-effector function (B) profiling of polyclonal antibodies against the indicated antigens. The cell color indicates the odds ratio within the group according to the key below the graph. Asterisks indicate Benjamini-Hochberg-adjusted p-values for Mann-Whitney U tests (•p < 0.1; *p < 0.05). N.T, not tested.

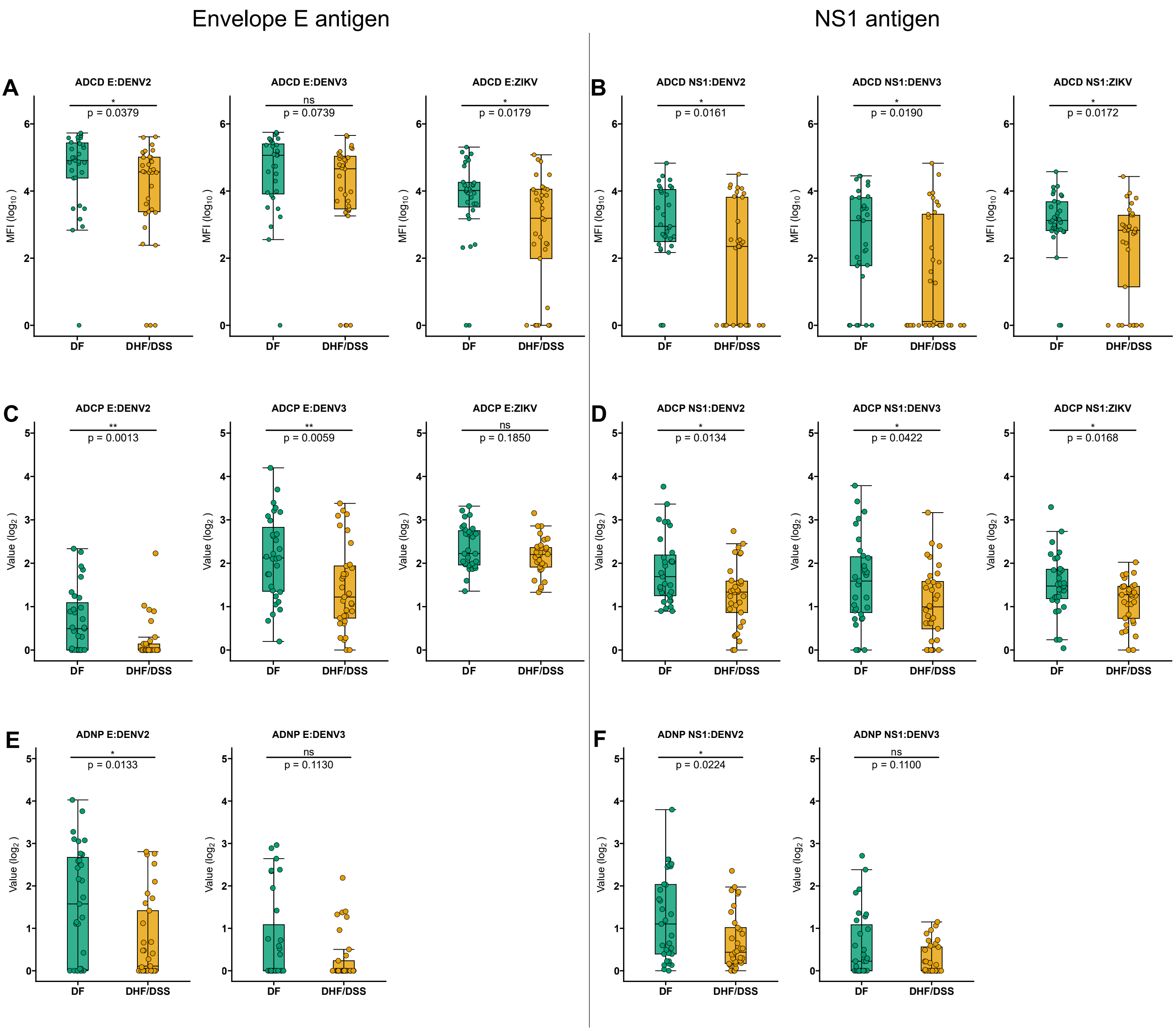


**Fig. S4. ADCD, ADCP and ADNP profiles in pre-infection samples from participants who experienced subsequent DF vs DHF/DSS.** (A,C,E) Envelope (E) antigen. (B,D,F) NS1 antigen. (A,B) ADCD with E, left panel; ADCD with NS1, right panel. (C,D) ADCP with E, left panel; ADCP with NS1, right panel. (E,F) ADNP with E, left panel; ADNP with NS1, right panel. Shown are median (middle line), 25th to 75th percentile (box), and 5th to 95th percentile (whiskers) as well as the raw data (points). Asterisks indicate Benjamini-Hochberg adjusted p-values for Mann-Whitney U tests (*p < 0.05, **p < 0.01, and ns, non-significant).

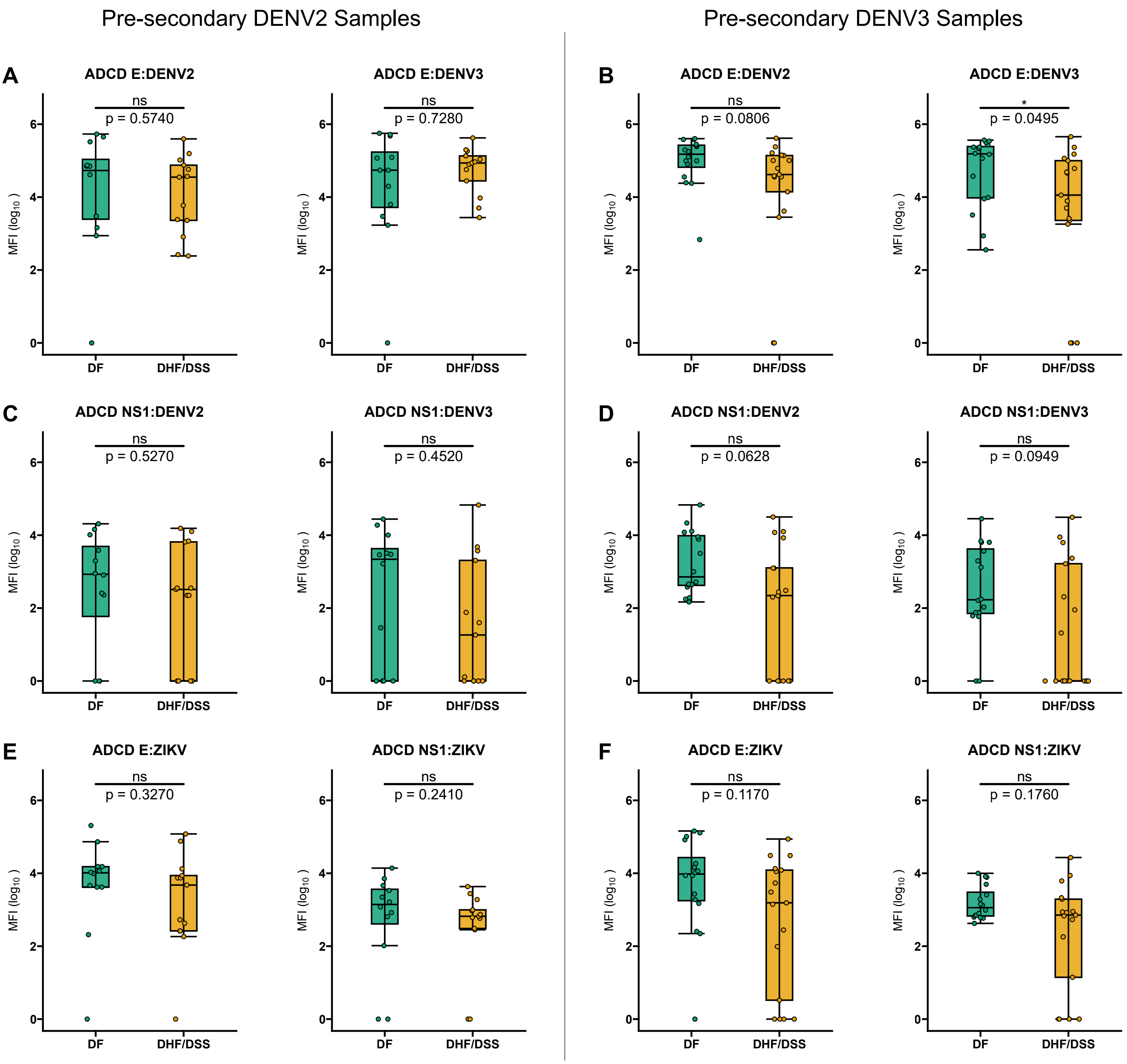


**Fig. S5. ADCD profiles in pre-infection samples from participants who experienced subsequent DF vs DHF/DSS DENV stratified by incoming serotype.** (A,C,E) Pre-secondary DENV2 samples. (B,D,F) Pre-secondary DENV3 samples. (A,B) ADCD with DENV2 envelope (E), left panel; ADCD with DENV3 E, right panel. (C,D) ADCD DENV2 with NS1, left panel; ADCD with DENV3 NS1, right panel. (E,F) ADCD with ZIKV E, left panel; ADCD with ZIKV NS1, right panel. Shown are median MFI (middle line), 25$^{th}$ to 75$^{th}$ percentile (box), and 5$^{th}$ to 95$^{th}$ percentile (whiskers) as well as the raw data (points). Asterisks indicate Benjamini Hochberg adjusted p-values for Mann-Whitney U tests (*p < 0.05, and ns, non-significant).

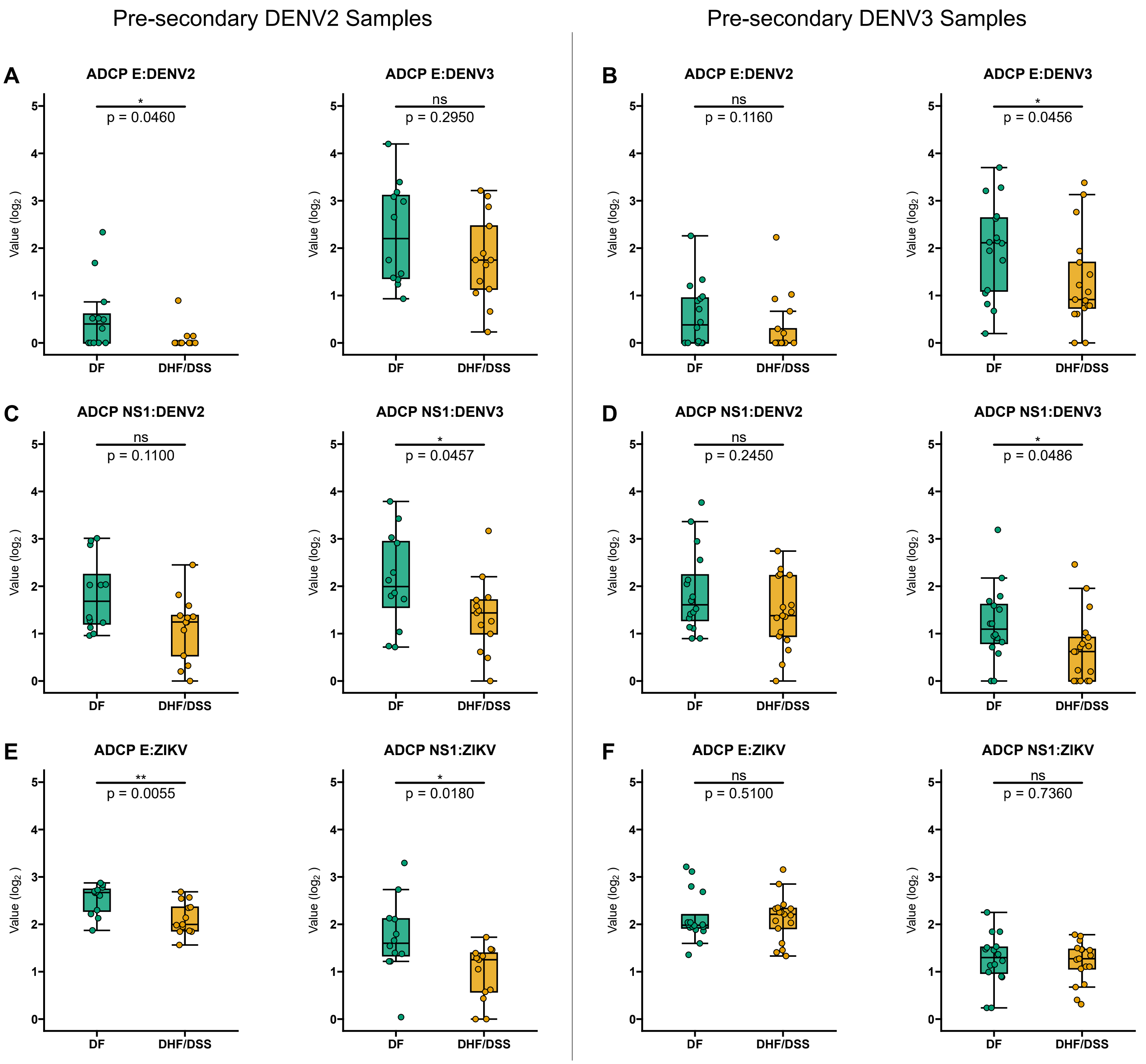


**Fig. S6. ADCP profiles in pre-infection samples from participants who experienced subsequent DF vs DHF/DSS stratified by incoming serotype.** (A,C,E) Pre-secondary DENV2 samples. (B,D,F) Pre-secondary DENV3 samples. (A,B) ADCP with DENV2 envelope (E), left panel; ADCP with DENV3 E, right panel. (C,D) ADCP DENV2 with NS1, left panel; ADCP with DENV3 NS1, right panel. (E,F) ADCP with ZIKV E, left panel; ADCP with ZIKV NS1, right panel. Shown are median (middle line), 25$^{th}$ to 75$^{th}$ percentile (box), and 5$^{th}$ to 95$^{th}$ percentile (whiskers) as well as the raw data (points). Asterisks indicate Benjamini Hochberg adjusted p-values for Mann-Whitney U tests (*p < 0.05, and ns, non-significant).

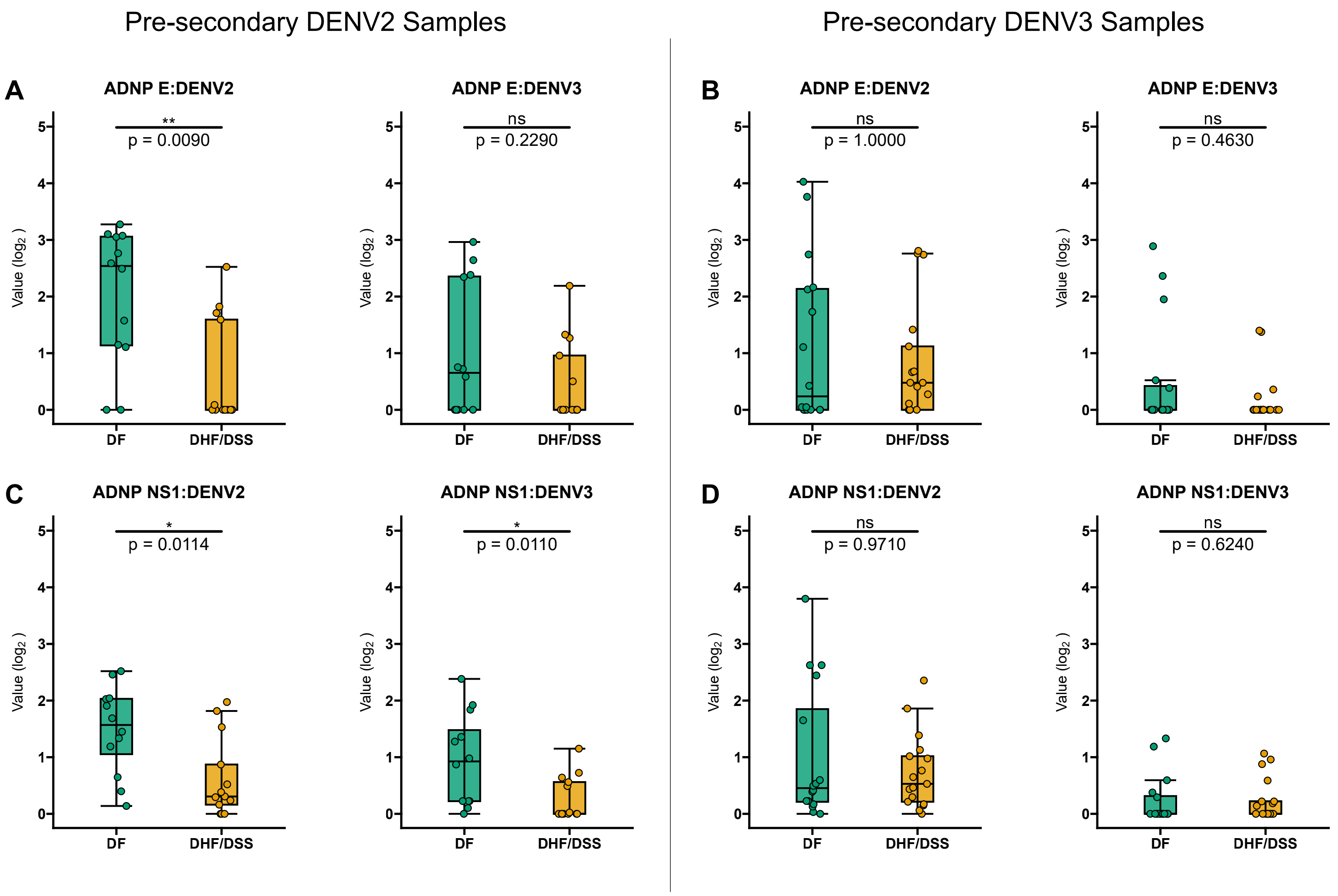


**Fig. S7. ADNP profiles in pre-infection samples from participants who experienced subsequent DF vs DHF/DSS stratified by incoming serotype.** (A,C,E) Pre-secondary DENV2 samples. (B,D,F) Pre-secondary DENV3 samples. (A,B) ADNP with DENV2 envelope (E), left panel; ADNP with DENV3 E, right panel. (C,D) ADNP with DENV2 NS1, left panel; ADNP with DENV3 NS1, right panel. Shown are median (middle line), 25$^{th}$ to 75$^{th}$ percentile (box), and 5$^{th}$ to 95$^{th}$ percentile (whiskers) as well as the raw data (points). Asterisks indicate Benjamini Hochberg adjusted p-values for Mann-Whitney U tests (*$p < 0.05$, **$p < 0.01$, and ns, non-significant).

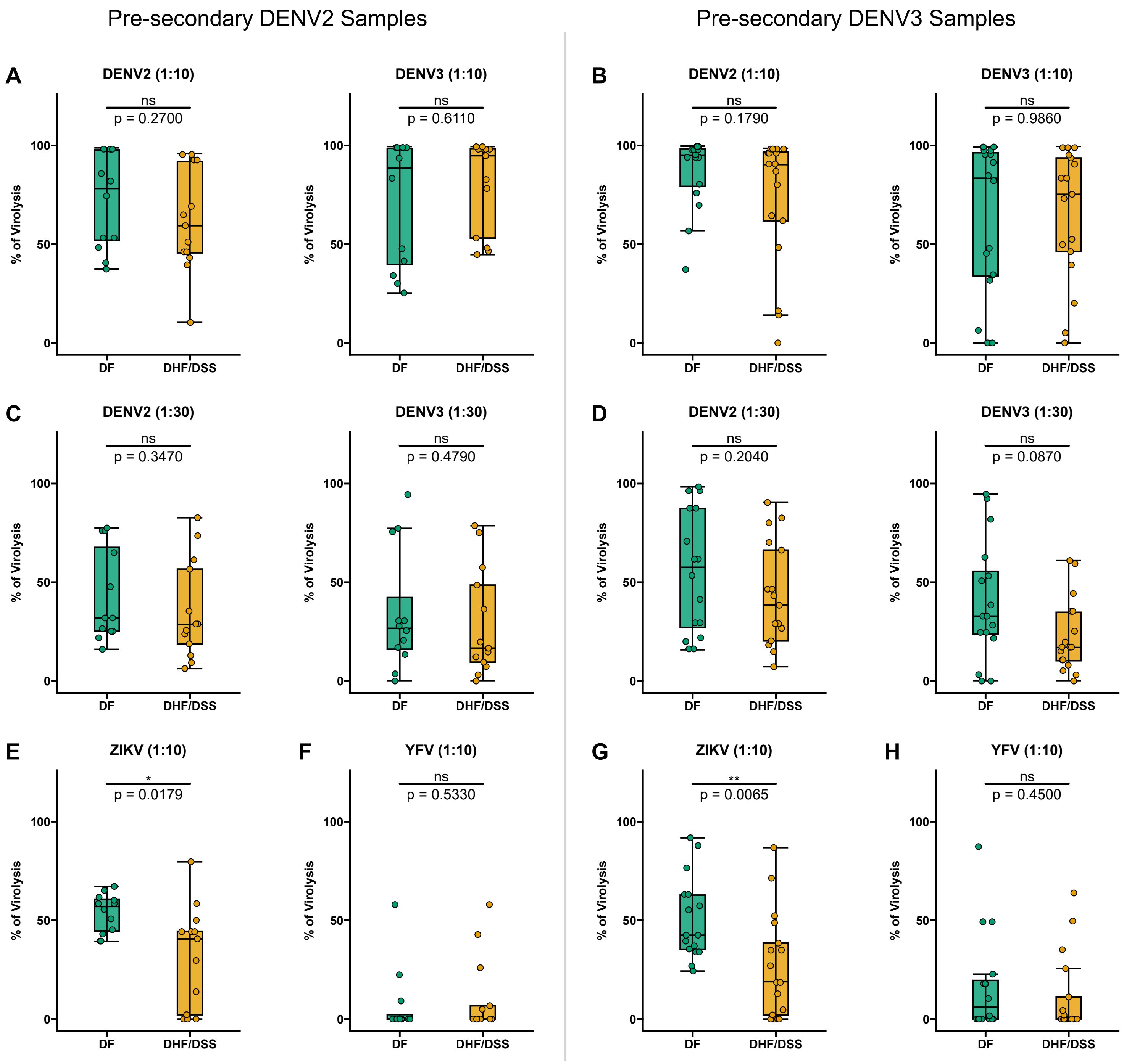


**Fig. S8. Virolysis profiles in pre-infection samples from participants who experienced subsequent DF vs DHF/DSS stratified by incoming serotype.** (A,C,E,F) Pre-secondary DENV2 samples. (B,D,G,H) Pre-secondary DENV3 samples. (A,B) DENV2, left panel; DENV3, right panel. (C,D) DENV2, left panel; DENV3, right panel. (E,F,G,H) ZIKV, left panel; ,YFV, right panel. 1:10 and 1:30 represent the dilution factor of the plasma for the assay. Shown are median % of virolysis (middle line), $25^{th}$ to $75^{th}$ percentile (box), and $5^{th}$ to $95^{th}$ percentile (whiskers) as well as the raw data (points). Asterisks indicate Benjamini Hochberg adjusted p-values for Mann-Whitney U tests (*p < 0.05, **p < 0.01, and ns, non-significant).
